# Derivation and Validation of a Point-based Forecasting Tool for SARS-CoV-2 Critical Care Occupancy

**DOI:** 10.1101/2025.01.21.25320912

**Authors:** Alicia A. Grima, Clara Eunyoung Lee, Ashleigh R. Tuite, Natalie J. Wilson, Alison Simmons, David N. Fisman

**Affiliations:** From the Dalla Lana School of Public Health, University of Toronto

## Abstract

**Background:** The requirement for critical care in even a modest fraction of SARS-CoV-2 infected individuals made ICU resources an important societal chokepoint during the recent pandemic. We developed a simple regression-based point score in 2020 based on an objective of forecasting critical care occupancy in the Canadian province of Ontario based on mean age of cases, case numbers, and testing volume. Evolution of the pandemic (variants of concern, vaccination) led us to re-assess and re-calibrate our earlier work, with inclusion of information vaccination which became widespread in 2021.

**Methods:** We obtained complete provincial SARS-CoV-2 case, testing, and vaccination data for the period from March 2020 to September 2022, with data subdivided into 6 major “waves”, following the approach applied by other Canadian investigators. Our initial model was fit only using the first two “wild type” SARS-CoV-2 waves; an updated model included wave 3 (N501Y+ variants). Our model was validated by comparing model projections to waves not used for model fitting; validation model fits were evaluated with Spearman’s rho; counterfactuals without vaccination were modeled to impute fraction of critical care admissions prevented with vaccination. Costing was based on published economic estimates.

**Results:** Our initial model (fit to waves 1 and 2) was well calibrated (rho 0.85) but predictive validity was modest (rho 0.46). Predictive validity improved in models fit to the first 3 pandemic waves without vaccination (rho 0.60) or with vaccination (rho 0.68) (P for inclusion of vaccination 0.013 by Likelihood Ratio Test). Prevented fraction of ICU admissions attributable to vaccination was 144% (22017 admissions expected vs. 9020 observed); based on published estimates of ICU admission cost for SARS-CoV-2 the 12977 admissions averted $2.9 (CDN) billion in economic costs, in contrast to the $3 billion total cost of the vaccination program.

**Conclusions:** Simple time series regression incorporating case and testing characteristics continues to be useful as a tool for forecasting critical care occupancy due to SARS-CoV-2 but early pandemic models need to be updated to capture the preventive effects of widespread vaccination. The economic benefit of vaccination for prevention of critical care resource consumption during the pandemic is substantial, achieving near cost neutrality with the province’s entire vaccination program.

## Background

A signature feature of the SARS-CoV-2 pandemic has been its impact on critical care capacity (1–5). While the heterogeneity of SARS-CoV-2 infection is remarkable, ranging from true asymptomatic infection to critical illness (6), requirement for critical care in even a modest fraction of infected individuals makes critical care resources an important societal chokepoint when communities experience high attack rates (1, 3–5). Saturation of critical care capacity can result in catastrophic care outcomes for those infected with SARS-CoV-2 and for those requiring critical care for other reasons (5).

Over the course of the year 2020, we noted that critical care admissions in the Canadian province of Ontario were not simply a function of case volume, but also the test volume upon which a given case count was based, and well as the risk level (captured by average case age) of those with reported infections. Mean case age, as well as case and test volumes, could be transformed into a simple regression-based point score that could predict lagged ICU admissions (7). However, subsequent to the year 2020, several important changes occurred in the dynamics of the pandemic: vaccination against SARS-CoV-2 became widely available in Canada, and novel SARS-CoV-2 variants of concern emerged, with initial N501Y-positive variants (Alpha, Beta, Gamma) becoming dominant in spring 2021 (8, 9), only to be replaced by the Delta variant in summer and autumn of 2021 (10), which was in turn replaced by Omicron variants in December 2021 (11).

## Objectives

Our initial objective was to create a simple, point-based scores that could allow health systems to plan for increased demand for critical care services in a manner that accounted for the lag between SARS-CoV-2 case occurrence and demand for critical care beds. With the passage of time and the evolution of SARS-CoV-2 epidemiology from that of a pandemic to that of a hyper-endemic disease, we sought to reassess the validity of our earlier point score, to update the score to account for the effects of vaccination, and (given the observed effects of vaccination in our model) to quantify the impact of vaccination on critical care usage by modeling counterfactual scenarios where vaccination did not occur.

## Methods

This study was approved by the Research Ethics Board of the University of Toronto (protocols #39239, #39253, and #44787). We obtained COVID-related ICU admissions, newly reported case counts, average weekly case age, and testing volumes, and vaccination coverage for the province of Ontario from March 17, 2020 (when epidemic spread of SARS-CoV-2 was clearly underway in the province) to September 4, 2022, when data availability ended, using Ontario’s Case and Contact Management (CCM) System, Ontario Laboratory Information System (OLIS), and the COVAX vaccine information system as described elsewhere (12, 13).

We constructed negative binomial regression models that predicted weekly ICU admissions based on average case age, log_10_(weekly test volumes) and log_10_(weekly case counts), and cumulative provincial vaccination levels (defined as total administered vaccine doses divided by population) at 2-week lags based on an average lag between symptom onset and ICU admission of 9.5 days (IQR 5 to 11 days).

Population denominators were obtained from Statistics Canada (14). We divided our time series into 6 SARS-CoV-2 “waves”, using the approach described by Mitchell et al.(15), with the first two pandemic waves caused predominantly by wild type virus, the third caused by N501Y+ variants, the fourth caused by the Delta variant, and the final two waves caused by Omicron variants (15).

### Regression Models

Our initial predictive model was fit only to the first 2 (wild type virus) waves of the pandemic. This model was validated by comparing model predicted weekly critical care admissions to observed admissions over the subsequent four waves. As the model had limited predictive validity, we refit our earlier model to the first 3 pandemic waves, both without and with a vaccination term. Predictive validity was assessed by evaluating correlation between model predictions and observed admissions using Spearman’s ρ. Model fits with and without vaccination were compared using the Likelihood Ratio test.

### Simplified Point Scores

For ease of application, we converted our regression models to simple point scores by dividing model coefficients by the value of the smallest coefficient (which itself received a point score of 1). As point scores were based on log-linear negative binomial models, point scores could be converted into expected critical care occupancy by multiplying point scores by the weekly value of covariates (log_10_(cases), log_10_(tests), average age of cases, and for models including cumulative vaccination, vaccine doses per capita. These weighted point scores were then summed, multiplied by the value of the smallest model coefficient, and exponentiated to obtain predicted critical care admissions. For example, if the scores associated with log_10_(weekly cases), mean weekly case age, and log_10_(weekly tests) were 50, -1, and -60, and the point score for the model intercept as 200 points, one would obtain the score S = 50(log_10_(weekly cases)) – 1 (mean age) – 60 (log_10_(weekly tests)) + 200. If the multiplier (based on the coefficient for age) were 0.05, predicted critical care admissions e^(0.05 x S)^.

### Prediction of Occupancy

The rate of critical care admissions is only one of two factors that determine occupancy, with the second being mean length of stay of critically ill individuals.

While mean length of stay would ideally be estimated using survival analysis methods, a high proportion of individuals in the CCM dataset who were admitted to critical care settings lacked either a live discharge date or death date. As such, we estimated length of stay by fitting a simple difference equation model to occupancy data, such that:

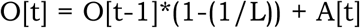

Here O[t] is mean weekly critical care occupancy at some time t, L is mean length of stay in weeks, and A[t] is admissions per week. Mean weekly SARS-CoV-2 occupancy estimates for Ontario were obtained from the Canadian Institutes for Health Information. We subsequently evaluated the predictive validity of our point score for critical care occupancy using a revised prediction model incorporating both admissions point scores, and length of stay estimates derived through fitting. We generated credible intervals for length of stay by treating both occupancy and admission time series as Poisson-random variates, and performing 1000 fits to these time series, with the 95% credible interval based on the range of the 950 best-fit values.

### Counterfactuals and Economic Benefit of Vaccination

The improved fit of our models to observed data when population-level vaccination coverage was incorporated may represent an index of critical care admissions prevented through widespread use of SARS-CoV-2 vaccines. We used our models to estimate the prevented fraction of SARS-CoV-2 critical care attributable to vaccination by generating model predictions with the coefficient for vaccination set to zero, as described by Zhou (16). Prevented fraction was obtained by comparing model predicted critical care admissions with and without vaccination.

We estimated the economic value of critical care admissions averted based on estimates of mean critical care costs per admission of $47,883 ($CDN) generated by the Canadian Institute for Health Information (CIHI) (17). This cost was applied to all critical care admissions. We estimated a critical care case fatality of 35.4% based on CCM data, consistent with data published by CIHI (17). For survivors, we applied an additional excess cost associated with attributable medical care in the year after critical care discharge; Sander (18) and colleagues estimated an excess cost of $114,945 for the year after SARS-CoV-2 diagnosis in individuals who survived critical care admission; we assigned the difference between this cost and critical care admission costs ($57,794) to survivors. For those who died, we estimated the mean quality-adjusted life years (QALY) lost because of death in critical care, with QALY valued at $30,000 as described elsewhere (19). QALY losses per fatality are dependent on the age at which individuals die; we calculated a weighted average QALY loss of 12.57 QALY per critical care fatality based on QALY weights from Kirwin et al. (20) (**Supplementary Appendix**). We did not estimate QALY losses for individuals who survived critical care admission. We performed sensitivity analyses in which we excluded CIHI estimates of critical care costs and assigned the full excess cost of $114,945 to critical care survivors and $91,091 for critical care admitted individuals who died, based on the estimates of Sander et al. (18). Input data for economic analyses are presented in **Table 2**.

## Results

Weekly SARS-CoV-2-related critical care admissions, critical care occupancy, reported cases, tests per capita, mean age of cases, and SARS-CoV-2 vaccine uptake from March 2020 to September 2022 are presented in **Figure 1**. A model based on log10(cases), log10(tests) and average case age (“model 1”), fitted to the first two pandemic waves was well calibrated, but had modest predictive validity (Spearman’s ρ 0.46 for the last four pandemic waves) (**Table 1** and **Supplementary Appendix**).

**Figure 1.**
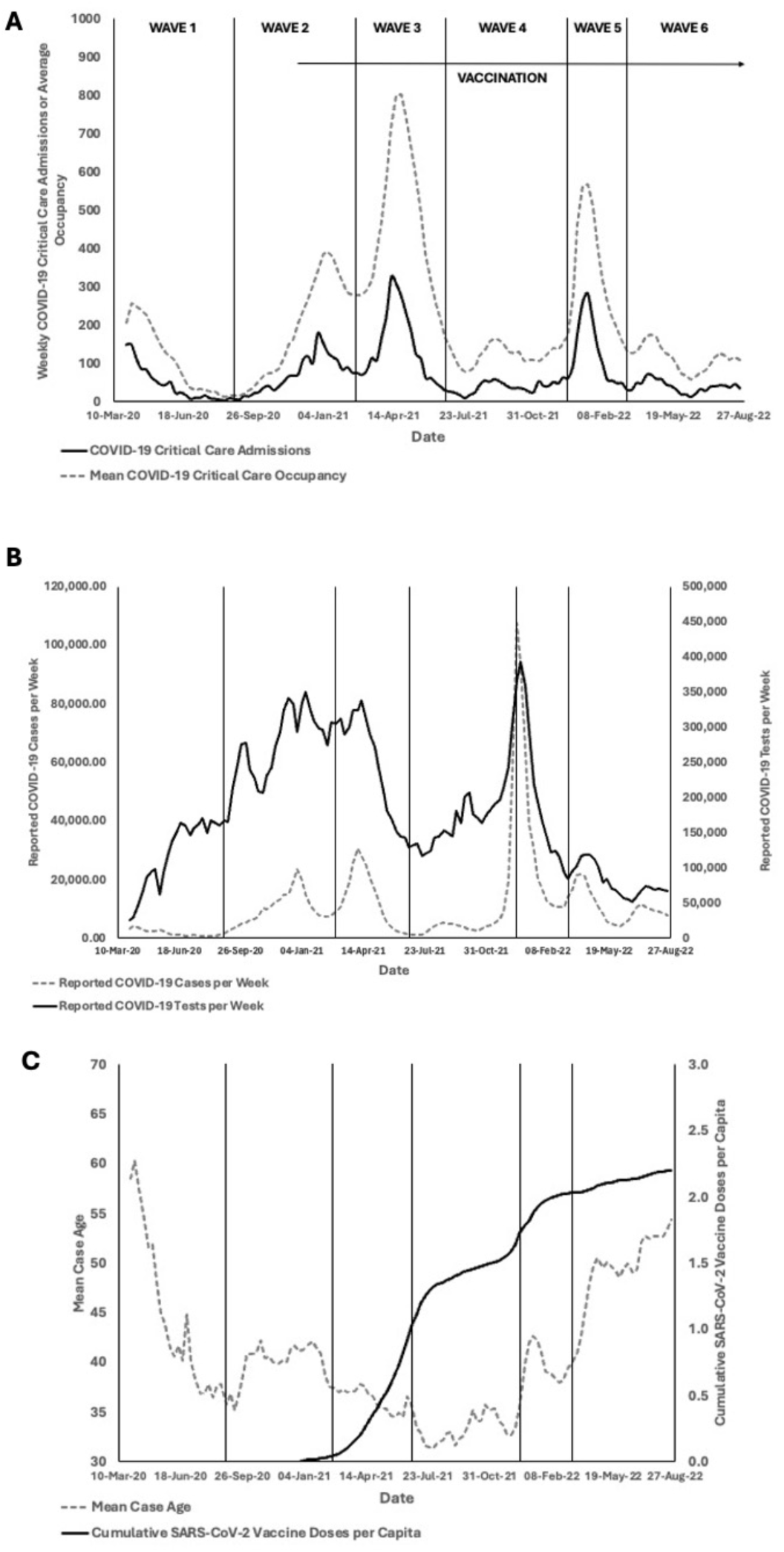
Epidemiology of the SARS-CoV-2 Pandemic in Ontario, Canada, from March 2020 to September 2022. Panel A (top) shows the number of COVID-19-related critical care admissions (black curve) and COVID-19 critical care occupancy (dashed curve) in the province. Successive waves are divided by vertical lines. Arrow denotes timing of the province’s vaccination campaign. Panel B (middle) shows the number of weekly reported COVID-19 cases (dashed curve, left Y-axis) and SARS-CoV-2 tests (black curve, right Y-axis) in the province; vertical lines again denote pandemic waves. Panel C (bottom) shows mean case age among reported COVID-19 cases (dashed curve, left Y-axis) and cumulative vaccination doses administered per person (black curve, right Y-axis). All panels: X-axis represents calendar date.

**Table 1.** Negative Binomial Models and Point Scores for Prediction of Weekly Critical Care COVID-19 Admissions.

|  | **Model 1** |  | **Model 2** |  | **Model 3** |  |
| --- | --- | --- | --- | --- | --- | --- |
|  | Coefficient (95% CI) | Points | Coefficient (95% CI) | Points | Coefficient (95% CI) | Points |
| **Intercept** | 11.569 (8.276 to 14.862) | 233 | 9.256 (6.235 to 12.277) | 299 | 11.355 (8.029 to 14.680) | 220 |
| **Log<sub>10</sub>(tests)** | -2.910 (-3.511 to -2.309) | -58 | -2.557 (-3.100 to -2.014) | -83 | -2.852 (3.427 to 2.277) | -55 |
| **Log<sub>10</sub>(cases)** | 2.661 (2.310 to 3.013) | 53 | 2.586 (2.288 to 2.885) | 83 | 2.685 (2.391 to 2.979) | 52 |
| **Mean age (years)** | -0.050 (-0.078 to -0.021) | -1 | -0.031 (-0.056 to -0.006) | -1 | -0.052 (-0.080 to -0.023) | -1 |
| **Vaccine doses per population** | --- | --- | --- | --- | -0.646 (-1.131 to -0.161) | 13 |
| **Multiplier** | --- | 0.050 |  | 0.031 |  | 0.052 |
| **Spearman's $\rho$** |  |  |  |  |  |  |
| **Derivation** | 0.850 | --- | 0.923 | --- | 0.899 | --- |
| **Validation** | 0.462 | --- | 0.603 | --- | 0.682 | --- |
| **P for log-rank test\*** |  |  | P = 0.013 |  |  |  |

**NOTE:** Model 1 fit to first two pandemic (wild type virus) waves; models 2 and 3 fit to first three pandemic waves without or with inclusion of vaccination, respectively.
\*Comparison of fit of model fit to first two pandemic waves without inclusion of vaccination to same model with vaccination included.

A model that used the same covariates as model 1 (“model 2”), but was fit to data from the first three, rather than two, pandemic waves had improved predictive validity (Spearman’s ρ 0.60 for last three pandemic waves (**Table 1**)). Predictive validity that was further improved by including vaccine doses per capita as an independent variable (“model 3”) (Spearman’s r 0.68) (**Table 1**). Graphical inspection demonstrated that model 3 predictions (which incorporated vaccine effects) fit observed critical care admissions better than those of model 2 (**Supplementary Appendix**), with a significant improvement in model fit with inclusion of vaccination (P = 0.013 by likelihood ratio test) (**Supplementary Appendix**).

Model derived point scores are presented in **Table 1**. Points for model intercept, average age, log10(cases) and log10(tests) were similar between model 1 (fit to the first two pandemic waves) and model 3 (which included vaccine doses per capita). Modifying the point score derived from model 1 by including the vaccine point score from model 3 resulted in predictions almost identical to those from the model 3 point score (**Figure 2**).

**Figure 2.**
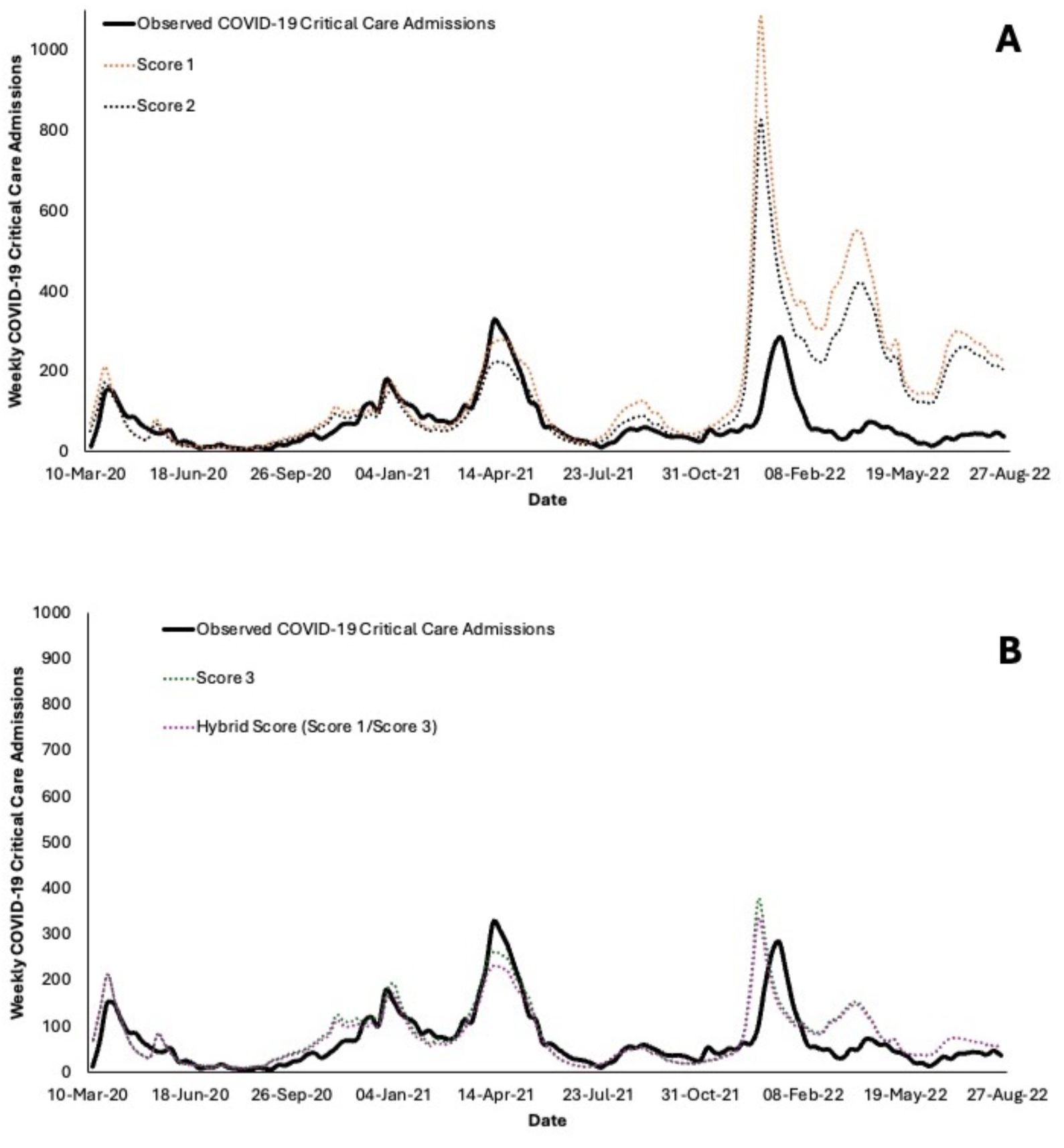
Predicted COVID-19 Critical Care Admissions Based on Point Scores. Top panel (A) shows observed critical care admissions per week (black curve), with predicted admissions based on point scores derived from models fitted to the first two pandemic waves (score 1, orange dashed curve) or first three pandemic waves (score 2, blue dashed curve). Bottom panel (B) shows improved fit for score 3 (blue dashed curve, based on a model fitted to the first three pandemic waves, but with the addition of cumulative vaccination) and for a hybrid score (score 1 with the addition of score 3-derived points for vaccination (red dashed curve).

We estimated length of stay through least squares fitting based on observed admissions and occupancy, with a mean length of stay estimate of 3.04 weeks (95% credible interval 2.96 to 3.13 weeks) providing excellent fit to data (**Figure 3**).

**Figure 3.**
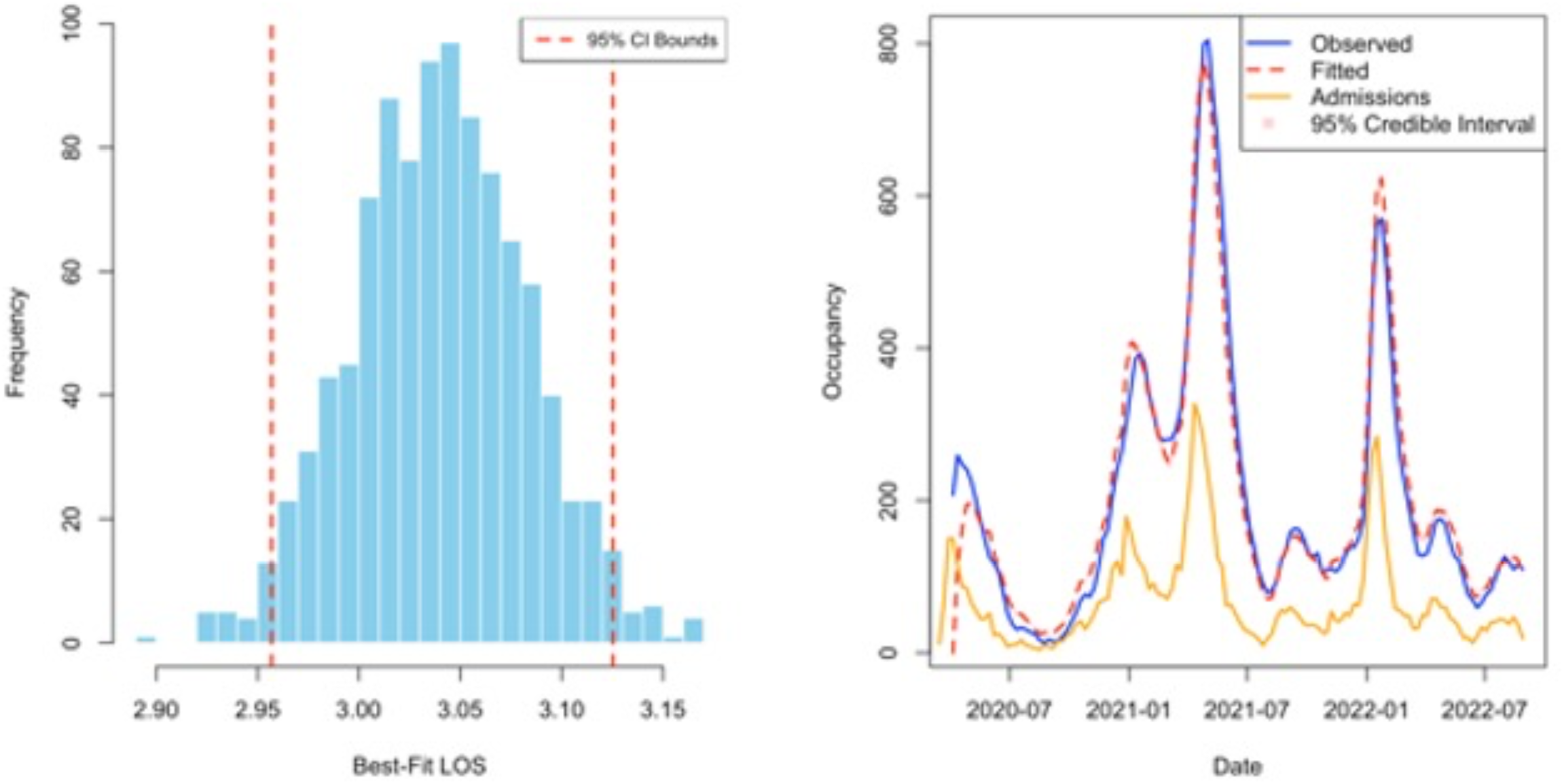
Estimated Critical Care Unit Length of Stay (LOS). Left panel shows histogram of best fit estimates for average critical care length of stay among individuals admitted to Ontario critical care units with COVID-19 (best fit 3.04 weeks; 95% credible interval 2.96 to 3.13). Right panel: comparison of estimated occupancy based of LOS estimates combined with observed weekly admissions to observed weekly critical care occupancy time series (observed data, blue curve; modeled data, dashed red curve). Pink shading represents 95% credible interval; orange curve represents observed weekly admissions.

Predicted occupancy, based on admissions from point score 3 or the hybrid (score 1/score 3) point score, combined with length of stay of 3 weeks, showed an excellent fit to observed occupancy data (**Figure 4**).

**Figure 4.**
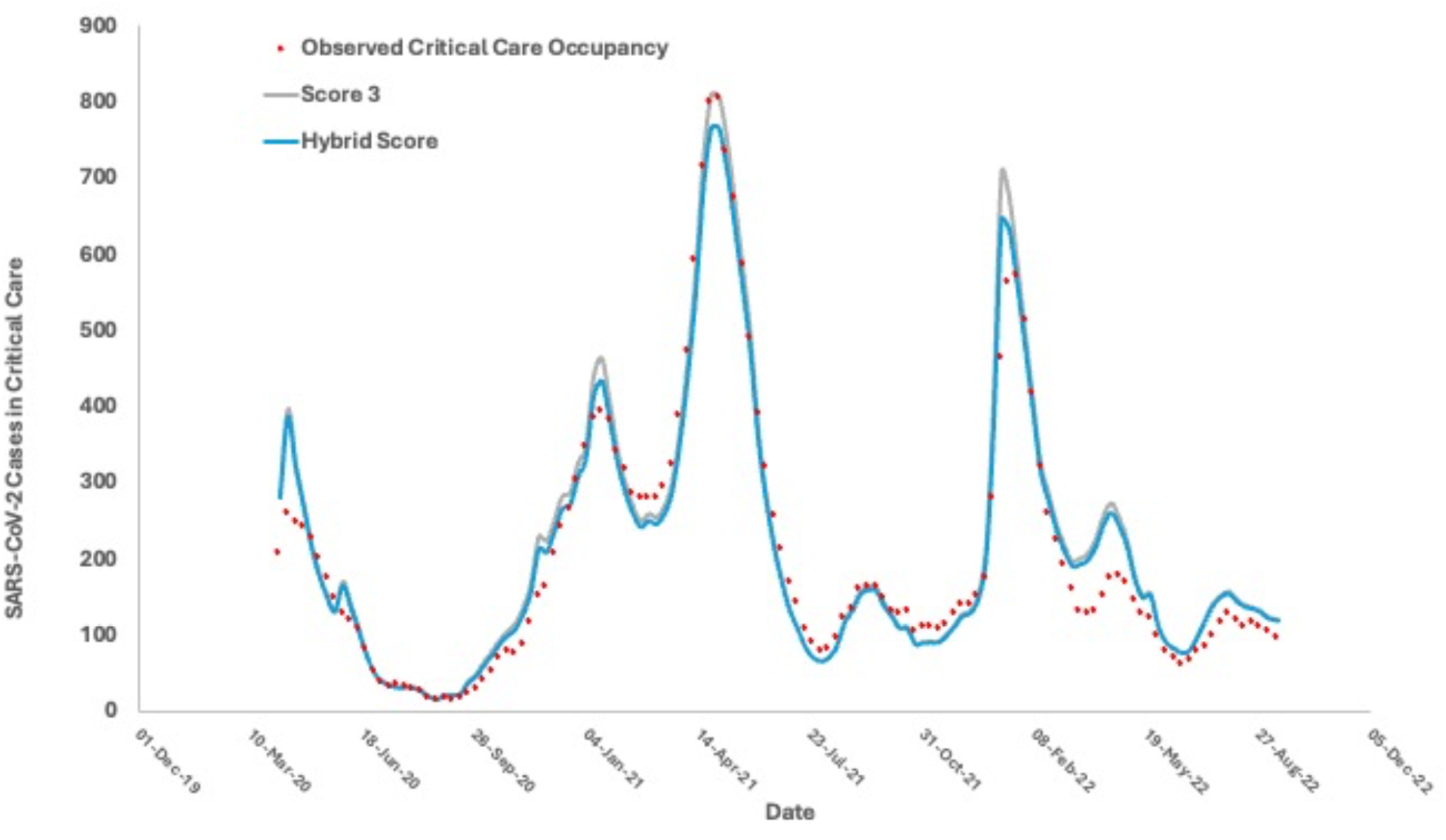
Predicted Critical Care Occupancy based on Model-Derived Point Scores. Predicted occupancy is based on prior week’s predicted occupancy, combined with admissions predicted using cases, tests, mean age, and vaccination coverage from two weeks prior. Score 3 is derived from model 3, a negative binomial model 3, with predictors including cases, tests, mean age and vaccination, fit to the first three pandemic waves. Hybrid score uses point scores for cases, tests and mean age from model 1 (fit to the first two pandemic waves) combined with vaccine points from score 3. Red points represent observed mean weekly critical care occupancy.

Prevented fraction of critical care admissions attributable to vaccination was 144% (22017 admissions expected vs. 9020 observed) (**Supplementary Appendix**); based on published estimates of ICU admission cost for SARS-CoV-2 the 12977 admissions averted $2.9 (CDN) billion in economic costs. Based on vaccine and programmatic cost estimates critical care-related costs averted through vaccination were approximately equivalent to the estimated cost of Ontario’s entire public vaccine program during the period under study ($3.0 billion). In sensitivity analyses that applied higher costs for critical care medical costs as well as subsequent attributable costs, critical care costs averted ($3.1 billion) exceeded vaccine program costs (**Table 2** and **Supplementary Appendix**).

**Table 2.** Costs Associated with Critical Care Admissions Averted and Vaccination Program.

| **Costs** | **Value** | **Reference** |
| --- | --- | --- |
| **Critical care costs averted** |  |  |
| **Critical care admissions averted** | 12997 | Present modeling |
| **Critical care case fatality** | 35.4% | CCM |
| **Critical care deaths averted** | 4549 | Present modeling |
| **Critical care admission** | \$47,883 | (17) |
| **QALY loss per death** | 12.57 | (20) and CCM |
| **Valuation per QALY** | \$30,000 | (19) |
| **One-year attributable excess costs in survivors** | \$114,945 | (18) |
| **One-year attributable excess costs in non-survivors** | \$ 91,091 | (18) |
| **Post-critical care costs** | \$57,794 | (17, 18) |
| **Total costs averted** | \$2,903,740,605 | |
| **Total costs averted (sensitivity analysis)** | \$3,094,323,784 | |
| **Vaccination costs** |  |  |
| **Vaccine cost per dose (\$)** | \$30 | (28) |
| **Vaccine administration costs per dose (\$)** | \$34 | (28) |
| **Other programmatic costs per dose (\$)** | \$27 | (28) |
| **Total doses administered** | 33,404,529 | COVAX |
| **Total vaccination program costs** | \$3,039,812,139 | |

| Net costs |  |  |
| --- | --- | --- |
| Net cost—base case | \$136,071,534 | |
| Net cost—sensitivity analysis | -\$54,511,645 | |

NOTES: QALY, quality-adjusted life years; CCM, Ontario Case Contact Management System; COVAX, Ontario COVID Vaccine Database.

## Discussion

The marked variability in severity of SARS-CoV-2 infection by age makes surveillance of the pandemic using only case counts problematic. We show here that future critical care burden can be accurately forecast if case counts are used in conjunction with estimates of test frequency, mean case age and crude measures of population immunity (in this case vaccine doses received per capita). Others have previously noted that correction for test frequency is necessary for accurate forecasting and estimation of infection fatality (21, 22), and we have also noted previously that variability in population age structure is a key determinant of SARS-CoV-2 epidemic severity (23, 24), while the apparent mean age of cases may again reflect the intensity of testing in different age groups (24). Here we show that these elements considered in conjunction can provide a simple, valid approach to near-term forecasting of critical care utilization. A similar approach might be applied to mortality forecasting or hospitalization.

While our earlier work appeared to accurately predict critical care admissions in early 2021, in our current analysis we find that that initial simple model ceased to be valid in predicting critical care admissions and overestimated critical care admissions in the last three pandemic waves we analyzed. However, the accuracy of our model seemed to be reestablished once we introduced cumulative vaccination coverage into our model, which resulted in accurate prediction of critical care admissions during the 4^th^ (Delta variant) pandemic wave, and also markedly improved fit to admissions in the 5^th^ and 6^th^ (Omicron variant) waves. Indeed, notwithstanding the fact that these latter waves were not used for model fitting, our model-derived prediction scores, combined with a length of stay estimate derived via fitting, led to an accurate projection of critical care occupancy in Ontario during the pandemic.

Our estimate of COVID-19 associated mean length of stay warrants comment. Our estimates of critical care admissions and occupancy come from two different data sources, but we found that these measures could be readily converted back and forth using a simple model with a single unknown (length of stay (LOS)), solved via fitting. Our value of 3.04 weeks (21.3 days) was far longer than the average critical care length of stay estimate of 10.3 days published by the Canadian Institutes for Health Information. Our decision to estimate LOS with model fitting reflected the large percentage of critical care-admitted individuals in the Ontario CCM dataset who were missing either a death date or a discharge date, which led us to suspect that such dates may not be recorded reliably, and indeed may be missing in an informative manner (25), which could in turn result in biased estimates of critical care LOS for COVID-19. The much longer average that we identified likely represents right-skewing of the length of stay distribution; such skewing is extremely important when modeling critical care resources as individuals with prolonged critical care stays reflect a factor that places the critical care system at risk of overflow.

Conceptually, our finding that case counts need to be presented in context of who (demographically) is being tested or infected (as reflected in a changing age distribution of cases) and how many individuals are tested (regardless of age) to adjust case counts for the intensity of surveillance, speaks to the degree to which testing activities drive perceived risk during outbreaks, epidemics and pandemics. Our finding that there is an inverse relationship between mean case age and critical care admissions may be counterintuitive, as severity of SARS-CoV-2 infection increased markedly with age. However, older case age may represent a greater intensity of testing in settings such as long-term care facilities, whose residents are unlikely to be admitted to critical care units. Unfortunately, neither test volumes nor mean case-age are not often reported or publicly available (though the latter can be approximated when cases are reported in discrete age groups). Our findings suggest that they represent an important data source that can be used to place crude reported case counts in context.

Incorporation of vaccination into our model markedly improved model fit to observed critical care admissions, suggesting that increasing coverage with SARS-CoV-2 vaccine blunted the severity of infections (26) such that (for a given level of testing and mean case age) the per-case risk of critical care admission was reduced over time. This allowed us to simulate a counterfactual scenario where vaccine was not available; we find that if vaccination was indeed driving the decline in per-case critical care risk, Ontario’s universal SARS-CoV-2 vaccination program effectively paid for itself, simply through prevention of critical care-related resource use.

While we were able to validate our approach using data from Ontario, limitations of this work include uncertainty around generalizability of this approach to other jurisdictions, and to larger and smaller geographic scales. Nonetheless, the apparent ability to accurately forecast critical care admissions and occupancy two weeks in advance (which can be achieved by recursive use of forecast occupancy data) may prove particularly useful to those charged with management of healthcare resources. While the SARS-CoV-2 pandemic represents less of a threat to populations and healthcare resources than it did at peak, the threat of other pandemic diseases which might impact critical care resources remains ever present (including the threat of influenza A H5NX at the time of writing) (27).

Given the aggregate nature of our time series data we cannot be certain that the vaccine effects we observe are causal, as opposed to other contemporaneous changes (increasing levels of exposure to the pathogen, or evolution of viral variants, for example). However, it is notable that improved prediction occurred across both Delta variant (wave 4) and Omicron variant (waves 5 and 6) waves, suggesting that the vaccine effects we observed are not due to widespread infection-derived immune experience, which only occurred in Ontario with Omicron emergence, or due to declining variant virulence, which appears to have been higher with the Delta variant than with other viral strains.

In summary, we present the derivation and validation of a simple point-based score to predict critical care resource demand from SARS-CoV-2. Our modeling approach could be used both for short-term forecasting focussed on SARS-CoV-2, and for forecasting related to other infectious threats such as influenza. The importance of incorporating vaccine coverage into our model highlights the degree to which a robust vaccination effort likely averted substantially greater demand for critical care resources in Ontario, in a manner that appears to have been approximately cost neutral.

## Supplementary Appendix

### Negative Binomial Model Fits

We fit negative binomial models to a time series of weekly critical care admissions in Ontario. Model 1 was used to predict admissions based on log10(cases), log10(tests) and mean age of cases two weeks prior. The model was fit to waves 1 and 2 of the pandemic, which occurred between March 17, 2020 and February 28, 2021. The time series used for fitting is represented by the gray curve; the vertical line represents the dividing line between wave 2 and wave 3. Weekly critical care admissions not used for model fitting are represented by the black curve. Model predictions (orange dashed line) were reasonable for wave 3 (N501Y+ variants) but overestimated critical care admissions for wave 4 (Delta variant, summer 2021), and for waves 5 and 6 (Omicron variants).

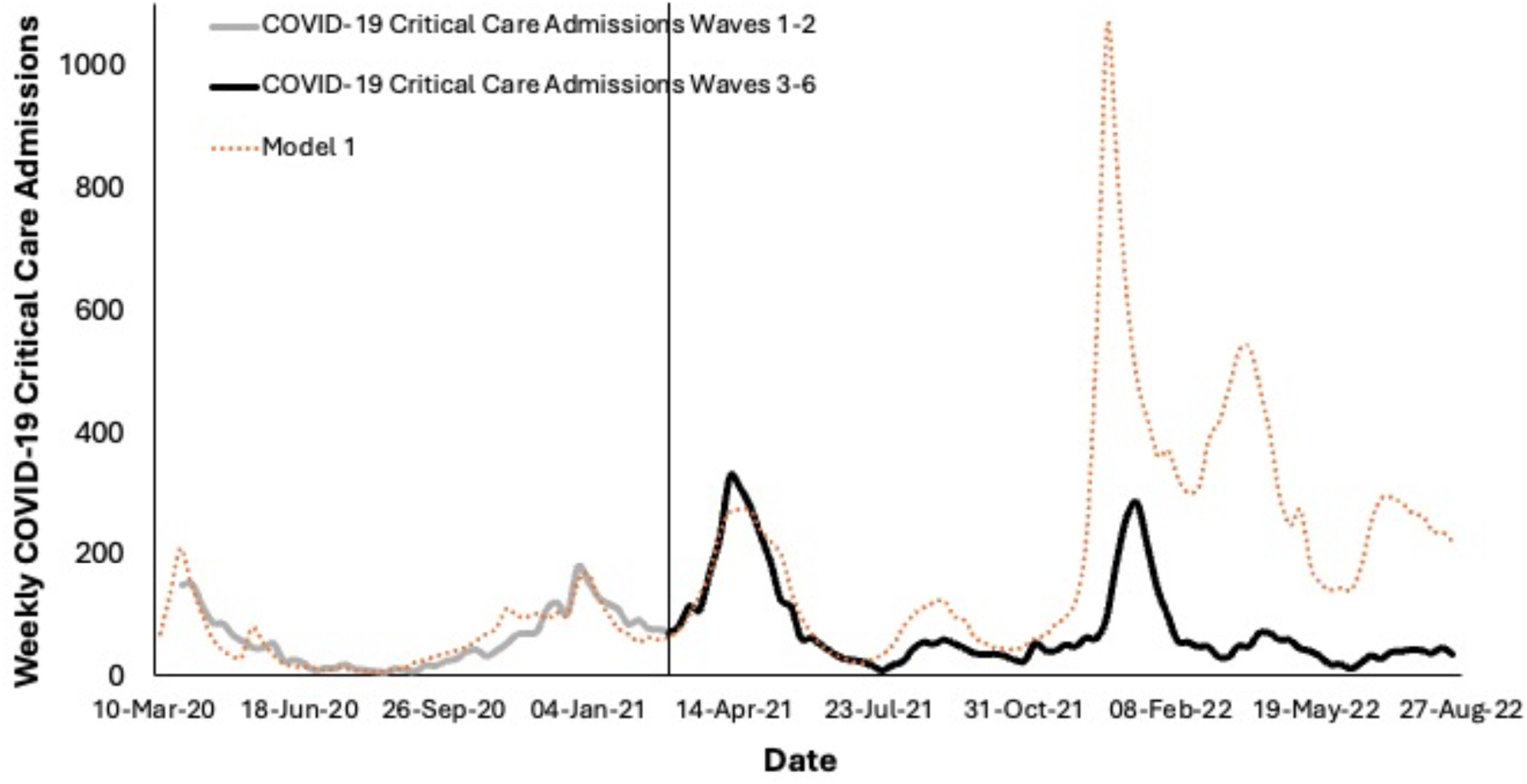

We subsequently fitted two additional models to waves 1-3, and used these to predict weekly critical care admissions. Model 2 was identical to model 1 aside from being fit to an additional pandemic wave. Model 3 differed from model 2 by incorporating cumulative administered SARS-CoV-2 vaccine doses per population as a covariate. The gray curve represents observed weekly critical care admissions used for fitting; the black curve represents admissions not used for model fitting. Model 2 is represented by the blue dashed curve and model 3 by the red dashed curve. As can be seen graphically, model fit was significantly improved with inclusion of vaccination (P = 0.013 by likelihood ratio test). Inclusion of vaccination resulted in excellent fit to wave 4 critical care admissions (July 2 to December 25, 2021), and markedly improved fit to waves 5 and 6 critical care admissions.

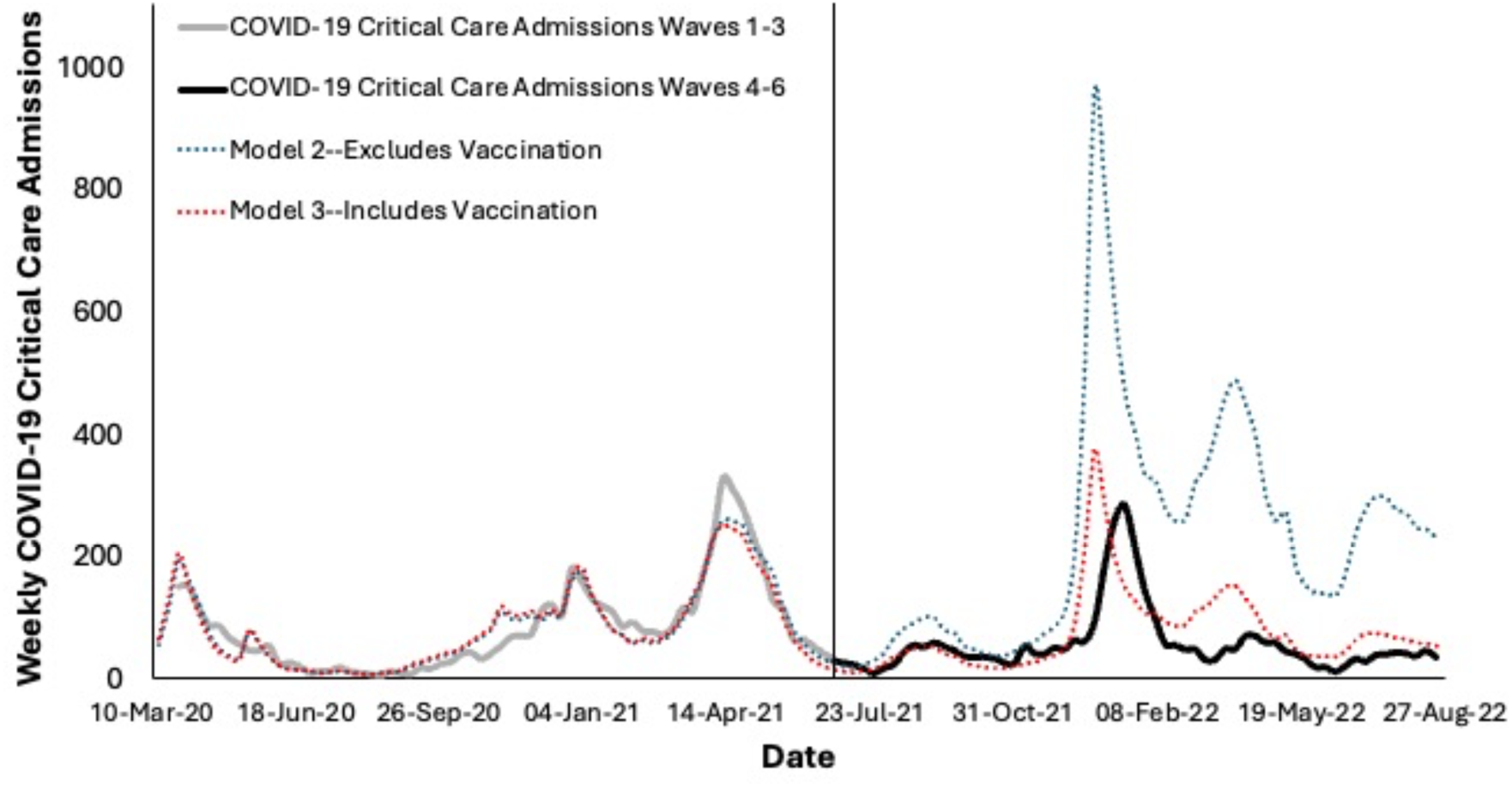

### Critical Care Admissions Averted via Vaccination

The marked improvement in model fit that occurred with the addition of cumulative vaccination identified the possibility that widespread population immunization against SARS-CoV-2 could have diminished the severity of SARS-CoV-2 infections (26) such that, for a given level of testing and mean case age, the per-case risk of critical care admission declined as vaccination coverage rose. We modeled a no-vaccination counterfactual by setting the coefficient for vaccination to zero in model 3 and comparing predicted cases in a model with vaccination to a model without vaccination. As noted in the results section of our paper, we estimated that the vaccination-prevented fraction of SARS-CoV-2 critical care admissions was 144% (12977 admissions averted) between March 2020 and August 2022. Predicted weekly critical care admissions with (blue curve) and without vaccination (tan curve) are presented below, with the yellow area between the two curves representing the prevented fraction of admissions.

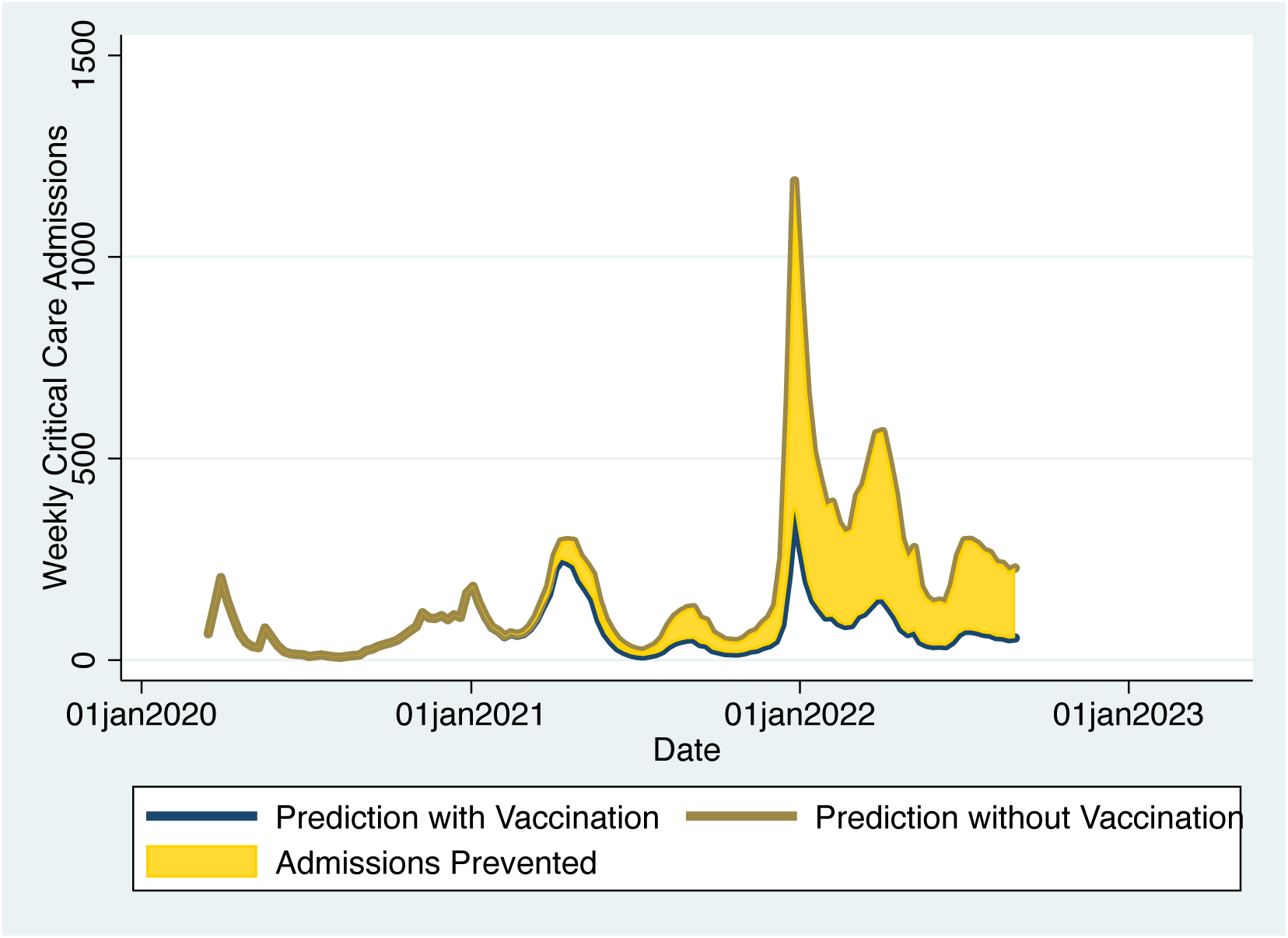

As noted in our Results section we estimated the economic value of critical care admissions averted by valuing each admission averted using the approach and costs outlined in **Table 2**, as well as estimating the monetary cost of Ontario’s public SARS-CoV-2 immunization program. The value of critical care admissions averted was approximately $2.9 billion ($CDN), which was slightly lower than the cost of the entire

Ontario public SARS-CoV-2 vaccine program ($3.0 billion). In a sensitivity analysis that used alternate data for valuation of critical care admissions, the value of admissions averted ($3.1 billion) exceeded the cost of the entire vaccine program. Estimated costs and components of costs are presented in the figure below.

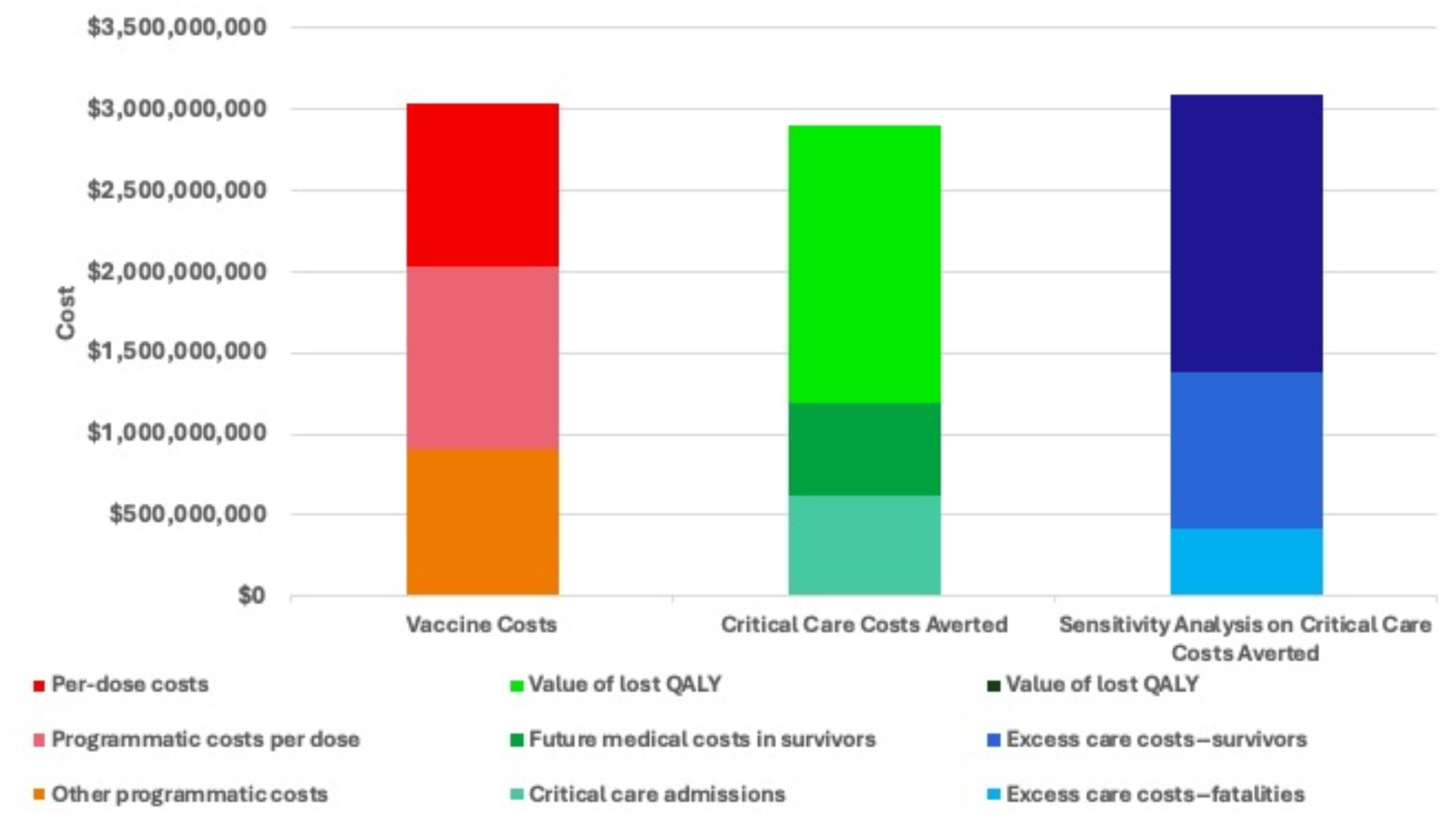

## Declarations

### Ethics approval and consent to participate

Approved by the University of Toronto Research Ethics Board, Protocols #39239, #39253 and #44787. Consent to participate: not applicable.

### Consent for publication

All authors have seen and reviewed the final submitted paper draft and consent to publication. Dr. Fisman takes responsibility for payment of any publication charges.

### Availability of data and material

Aggregate data and guidance on methods for reproduction of analyses are available from Dr. Fisman.

### Authors’ contributions

NW, AG, ART, CEL, AS and DNF contributed to the analysis and interpretation of data, drafted and/or substantially revised the work, have approved of the submitted version, and take responsibility for the submitted work. DNF and ART acquired necessary data and conceived of the work. DNF, ART and AG designed the study.

### Clinical Trial

Clinical trial number: Not applicable (not a clinical trial).

### Data Availability Statement

Aggregate data and guidance on methods for reproduction of analyses are available from Dr. Fisman.

## Data Availability

Aggregate data and guidance on methods for reproduction of analyses are available from Dr. Fisman.

